# Age-specific mortality and immunity patterns of SARS-CoV-2 infection in 45 countries

**DOI:** 10.1101/2020.08.24.20180851

**Authors:** Megan O’Driscoll, Gabriel Ribeiro Dos Santos, Lin Wang, Derek A.T. Cummings, Andrew S. Azman, Juliette Paireau, Arnaud Fontanet, Simon Cauchemez, Henrik Salje

**Affiliations:** Department of Genetics, University of Cambridge, Cambridge, UK; Mathematical Modelling of Infectious Diseases Unit, Institut Pasteur, UMR2000, CNRS, Paris, France; Department of Biology and Emerging Pathogens Institute, University of Florida, Florida, USA; Department of Epidemiology, Johns Hopkins Bloomberg School of Public Health, Baltimore, USA; Unit of Population Epidemiology, Division of Primary Care Medicine, Geneva University Hospitals, Geneva, Switzerland; Emerging Infectious Diseases Unit, Institut Pasteur, Paris, France; PACRI unit, Conservatoire National des Arts et Métiers, Paris, France

## Abstract

The number of COVID-19 deaths is often used as a key indicator of SARS-CoV-2 epidemic size. However, heterogeneous burdens in nursing homes and variable reporting of deaths in elderly individuals can hamper comparisons of deaths and the number of infections associated with them across countries. Using age-specific death data from 45 countries, we find that relative differences in the number of deaths by age amongst individuals aged <65 years old are highly consistent across locations. Combining these data with data from 15 seroprevalence surveys we demonstrate how age-specific infection fatality ratios (IFRs) can be used to reconstruct infected population proportions. We find notable heterogeneity in overall IFR estimates as suggested by individual serological studies and observe that for most European countries the reported number of deaths amongst ≥65s are significantly greater than expected, consistent with high infection attack rates experienced by nursing home populations in Europe. Age-specific COVID-19 death data in younger individuals can provide a robust indicator of population immunity.

---

As SARS-CoV-2 continues its rapid global spread, increased understanding of the underlying level of transmission and infection severity are crucial for guiding pandemic response. While the testing of COVID-19 cases is a vital public health tool, variability in surveillance capacities, case-definitions, testing indications, and health-seeking behaviour can cause difficulties in the interpretation of case data. Due to more complete reporting COVID-19 deaths are often seen as a more reliable indicator of epidemic size. If reliably reported, the number of COVID-19 deaths can be used to infer the total number of SARS-CoV-2 infections using estimates of the infection fatality ratio (IFR, the ratio of COVID-19 deaths to total SARS-CoV-2 infections). Estimates of the IFR derived from studies that carefully estimate the number of infected individuals in a particular setting can help make the link between deaths and total infections as well as refine estimates of the relative burden of mortality in different age groups^1^. While it is clear that infection severity increases significantly with increasing age^2,3^, there remain key unanswered questions as to the consistency of mortality patterns across countries. Underlying heterogeneities in the age structure of the population, or in the prevalence of comorbidities can contribute to differences in the levels of observed COVID-19 fatalities^4^. In addition, when looking at the total number of COVID-19 deaths, the level of transmission amongst the general population can be difficult to disentangle from large outbreaks in vulnerable populations such as nursing homes and other long-term care settings. Indeed for many countries, the SARS-CoV-2 pandemic has been characterized by a heavy burden in nursing home residents, with over 20% of all reported COVID-19 deaths occurring in nursing homes in countries such as Canada, Sweden and the United Kingdom^5^. In other countries, few COVID-19 deaths have been reported in nursing home settings such as in South Korea and Singapore due to successful epidemic control and/or shielding policies^5^. The reporting of COVID-19 deaths for older individuals can also be subject to inconsistencies across settings due to variable prevalence of comorbidities with which a COVID-19-associated death could be mistakenly attributed and varying practices of post-mortem testing for COVID-19. Age-specific COVID-19 death data can therefore provide valuable insights into the underlying nature of transmission, as the reporting of deaths amongst younger populations is likely to be more robust than that of elderly individuals.

In this context, simply comparing the total number of deaths across countries may provide a misleading representation of the underlying level of transmission. SARS-CoV-2 seroprevalence surveys, which estimate the number of people with detectable antibodies against the virus, provide valuable information on the proportion of the population that have ever experienced a SARS-CoV-2 infection at a given time-point^6–9^. These seroprevalence surveys, however, can be subject to a number of biases and variable performance of different serological assays can complicate the comparison of results across different studies^10^. Additionally, when exploring the relationship between the number of infections and deaths in an ongoing outbreak, adjustments for delays between infection and seroconversion, and seroconversion and death are critical to the interpretation of results. Here, we present a model framework that integrates age-specific COVID-19 death data from 45 countries with 15 national-level seroprevalence surveys, providing new insights into the consistency of infection fatality patterns across countries (Figure 1A). We use our model to produce ensemble IFR estimates by age and sex in a single harmonized framework as well as estimates of the proportion of the population infected in each country. Further, we use these estimates to reconstruct the expected number of deaths in older individuals (≥65 years), which we compare to reported deaths in each setting, highlighting heterogeneity in the burden of mortality amongst elderly individuals across countries.

**Figure 1.**
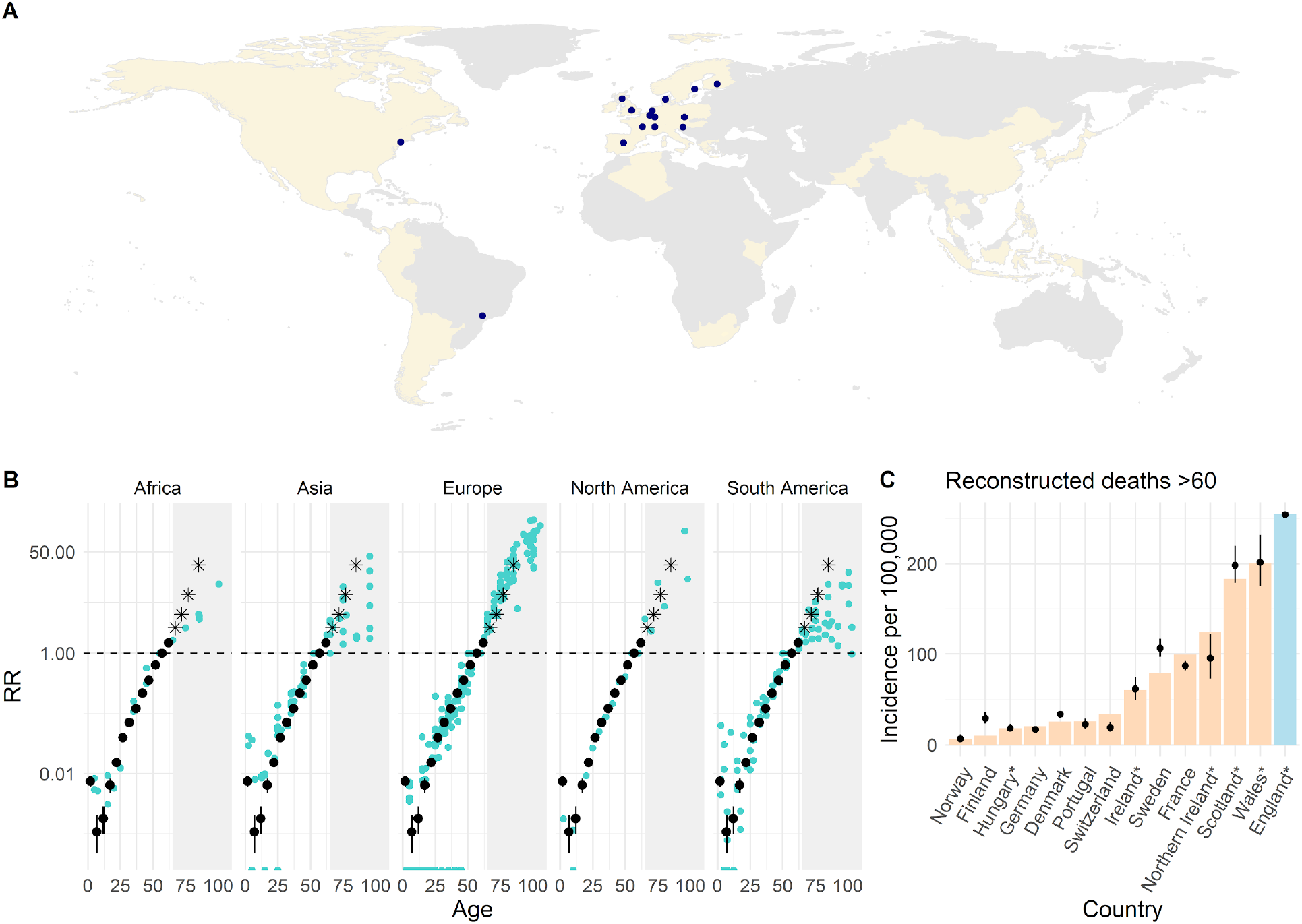
Patterns of COVID-19 mortality across settings. (A) Countries with age-specific death data (beige tiles) and locations with seroprevalence data (coloured points). (B) Estimated median and 95% credible interval (CrI) of the proportion of the population that have died in each age group, relative to the proportion that have died among 55-59 year olds in that country (black dots and lines), plotted on a log-linear scale. Coloured dots represent the country- and age-specific risks of COVID-19 death in the population relative to that of 55-59 year olds observed from reported death data, accounting for population age distributions (Supplementary Methods S1). All data points are plotted at the midpoint of the reported age group. The grey shaded areas highlight the relative risks of death by age for age groups ≥65, excluded from model fitting and black stars represent estimates inferred from England data only which are derived independent of nursing home deaths. (C) Comparing the reconstructed number of deaths with reported data for age-groups 60 or 65+ for a subset of countries where nursing home deaths could be excluded. Black dots and lines indicate the estimated median and 95% CrI; coloured bars show the reported incidence of non-nursing home deaths aged ≥60. Countries labelled with an asterisk * indicate where the number of deaths were reconstructed for ages 65+, to align with the reported age-groups for each country.

## Age-specific mortality patterns

Using population age structures and age-specific death data, we compare the number of deaths by age within each country, using the number of deaths in 60-65 year olds as the reference. We find a very consistent pattern in the relative risk of death by age for individuals <65 years old across countries and continents, with a strong log-linear relationship between age and risk of death for individuals 30-65 years old (Figure 1B, Supplementary Methods S1). The observed relative risk of death in older individuals appears substantially more heterogeneous across locations. Given the potential for important variability in mortality associated with nursing home outbreaks across countries, we first investigate mortality patterns specifically in the general population, using age-specific deaths ≥65 from England, where granularity of the data allows us to remove deaths in nursing home populations. We find that the log-linear relationship between age and risk of death continues into older age groups (Figure 1B). To assess the generalizability of data from England to other countries, we use these estimates to reconstruct the number of non-nursing home deaths reported in 13 other countries and find the predictions were consistent with the observed number of deaths in these countries (Figure 1C, Supplementary Methods S2).

**Figure 2.**
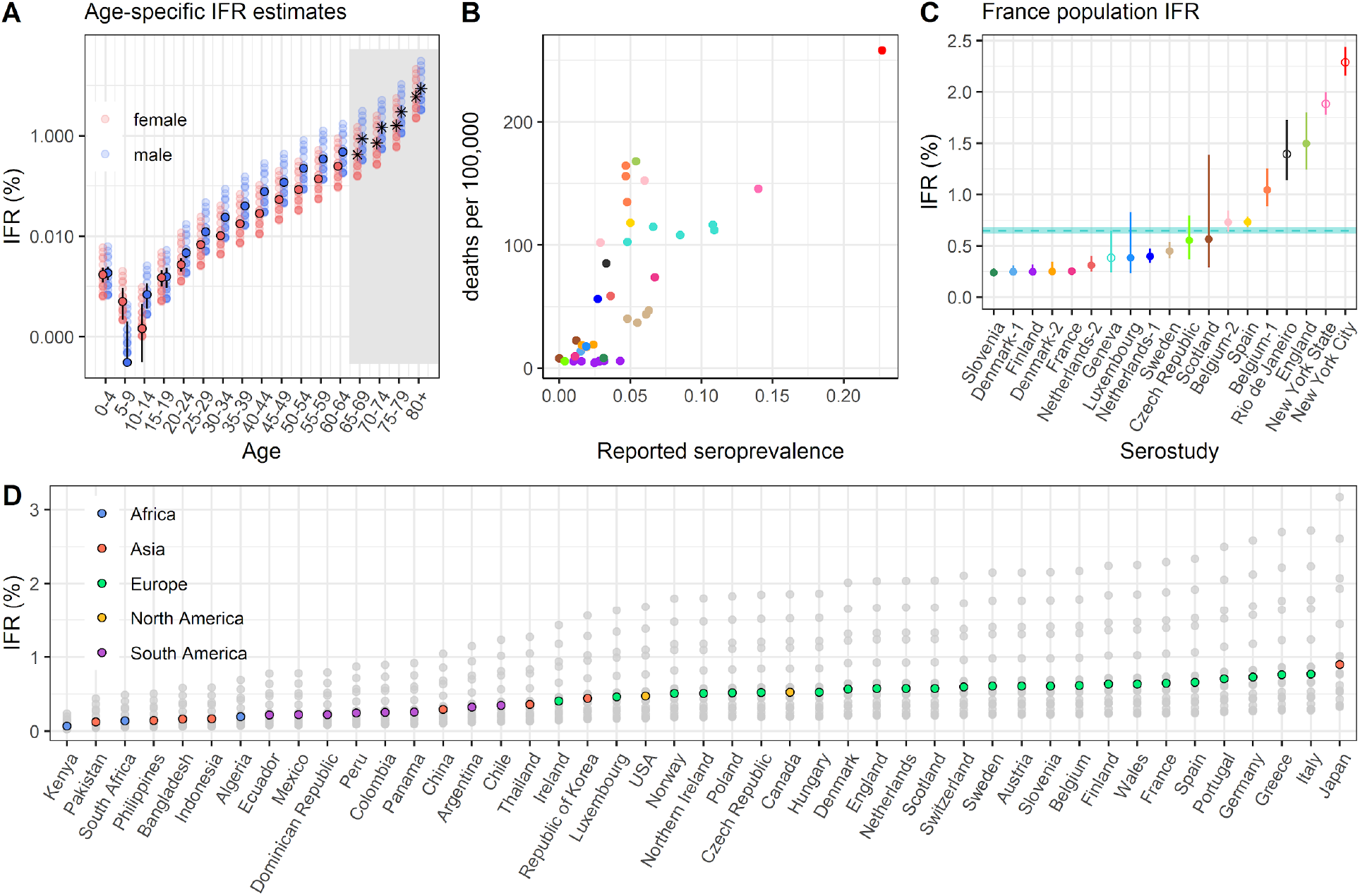
Infection fatality ratio (IFR) estimates. (A) Estimated median and 95% credible interval (CrI) of the IFR, stratified by age and sex and plotted on a log-linear scale. The IFR is estimated with the ensemble model (filled dots and black lines). Black stars on the right-hand side represent the estimated IFRs for age-groups ≥65, which were excluded from the fitting of the ensemble model. The coloured shaded dots represent median IFRs estimated from separately fitting to each individual serosurvey. (B) Relationship between the reported seroprevalence values and cumulative incidence of COVID-19 deaths 15 days after the end of seroprevalence sampling. Results from >1 serostudy were available for Belgium, Denmark, and Netherlands (Supplementary Table S1). (C) Using France as a reference country, population-weighted IFR estimates derived from fitting individual serological surveys in the model with points and lines indicating the median and 95% CrI. The blue dashed line and ribbon indicate the median and 95% CrI of the population-weighted IFR produced by the ensemble model. Hollow dots represent the estimates from subnational serological surveys that were excluded from the fitting of the ensemble model. (D) Median and 95% CrI of the population-weighted IFRs estimated by the ensemble model for each of the 45 countries, coloured by continent. Grey shaded dots represent the median estimates for each country, by fitting the model with each individual seroprevalence survey.

In order to translate relative risks of death by age to underlying IFR, we combine age-specific death data with 15 seroprevalence surveys, representing 12 of the 45 countries (2 different studies were each available for Belgium, Denmark and Netherlands, Supplementary Table S1). We use daily time-series of reported deaths to reconstruct the timing of infections and subsequent seroconversions. To limit biases that can be introduced by outbreaks in nursing home settings and the variable reporting practices of fatalities amongst individuals ≥65, we fit our model investigating the relationship between seroconversion and mortality exclusively to death data from those <65 years old. To infer IFRs in age groups ≥65 years, we use our estimates of the relative risk of death derived from England data only, without considering reported deaths from individual countries in these age groups. As our baseline model, we use an ensemble model where we include the results from all national-level seroprevalence studies together within a single framework. In addition, we consider separate models where we use the results of each serostudy individually to estimate IFRs in all locations, allowing us to investigate the consistency of estimates provided by different studies. As older individuals have fewer social contacts^11^ and are more likely to be isolated through shielding programmes we assume a baseline relative infection attack rate of 0.7 for individuals aged ≥65, relative to those <65, and assume equal infection attack rates across age groups <65 years. We find that age-specific IFRs estimated by the ensemble model range from <0.001% (95%CrI: 0-0.001) in those aged 5-9 years (ranging from 0-0.001% across individual national-level serostudies) to 7.27% (95%CrI: 6.91-7.66%) in those aged 80+ (ranging from 2.66-16.78% across individual national-level serostudies) (Figure 2A). A mean increase in IFR of 0.52% with each 5-year increase in age (95%CrI: 0.49-0.55%) was estimated for ages ≥10 years. We estimate that the risk of death given infection for men is significantly higher than that of women (Figure 2A) particularly in older individuals with ensemble IFR estimates of 8.62% for men aged 80+ (95%CrI: 8.19-9.07%) and 5.93% for women aged 80+ (95%CrI: 5.63-6.24%), consistent with previous findings^12,13^. Differences in ensemble IFR estimates by sex for age groups <20 years are less clear due to the small number of reported deaths in these age groups resulting in large uncertainty.

## Consistency of IFR estimates across seroprevalence surveys

Simple comparisons of the relationship between reported seroprevalence values and the cumulative incidence of COVID-19 deaths 15 days after the end of each seroprevalence survey, suggest large heterogeneity in the ratio of deaths to infections across settings (Figure 2B). We use our model framework to facilitate more robust comparisons of the IFR across settings, considering only age-specific deaths amongst <65 year olds. Using the country-specific demographic distributions (both age and sex) we estimate population-weighted IFRs for each country. Taking France as a reference population, the ensemble model estimates a population IFR of 0.65% for France (95% CrI: 0.62-0.68%) though we find notable heterogeneity in IFR estimates as suggested by individual seroprevalence studies, with a median range of 0.24-1.50% (Figure 2C). In particular, seroprevalence studies from England (1.50%, 95%CrI: 1.24-1.80%) and New York (1.88%, 95%CrI: 1.78-2.00%), both suggest a significantly higher IFR while studies in Slovenia (0.24%, 95%CrI: 0.22-0.28%), Denmark (0.25%, 95%CrI: 0.22-0.31%) and Finland (0.25%, 95%CrI: 0.22-0.32%) support a lower IFR than that of the ensemble model. Potential explanations for these differences include different prevalences of high-risk populations (e.g. individuals with comorbidities), differences in the methodology and representativeness of the seroprevalence studies, heterogeneities in the availability and quality of care or variations in the reporting of COVID-19 deaths. We find that studies conducted with blood bank sera (which do not include children and require individuals to be asymptomatic at the time of sample collection) gave similar results to studies in the general population (Supplementary Figure S5). Considering the demographic structures of each country, we find that population-weighted IFR estimates by the ensemble model are highest for countries with older populations such as Japan (0.90%, 95%CrI: 0.85-0.94%) and Italy (0.77%, 95%CrI: 0.73-0.81%), whilst the lowest IFRs among the 45 countries are for Kenya (0.07%, 95%CrI: 0.06-0.07%) and Pakistan (0.12%, 95%CrI: 0.12-0.13%) (Figure 2D).

Our ensemble model reproduces the reported seroprevalence values for the majority of studies including the dynamics of reported seroprevalence over time (Figure 3B). Of the 45 countries included in our analysis, representing 3.4 billion people, we estimate an average of 2.41% (95%CrI: 2.21-2.64%, individual serostudy range: 1.04-6.41%) of these populations had been infected by the 30th of May 2020 ranging from 0.07% (95%CrI: 0.05-0.09%, individual serostudy range: 0.03-0.18%) in Japan to 23.66% (95%CrI: 22.13-25.28%, individual serostudy range: 9.99-62.89%) in Ecuador. Consistent with other studies, these results indicate that the majority of countries are likely a long way from standard herd immunity thresholds at the national-level ^12,14,15^.

**Figure 3.**
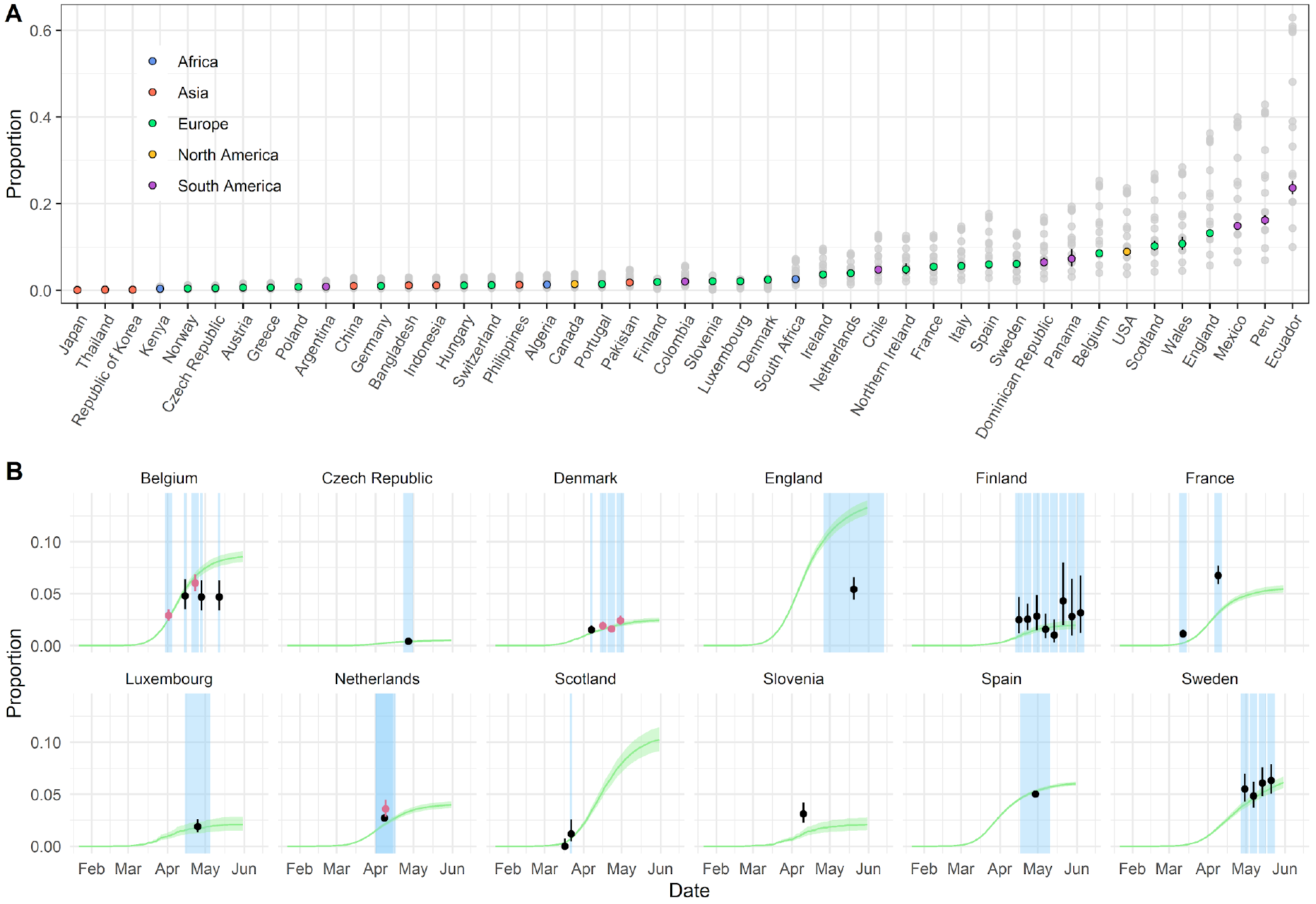
Infection attack rates. (A) Estimates of the infected population proportion for each country as of 30 May 2020. Grey shaded dots indicate the median estimates by fitting the model with each individual seroprevalence survey. Coloured dots and lines represent the median and 95% credible intervals (CrI) estimated by the ensemble model. (B) Proportion seropositive over time for each of the 12 countries with national-level seroprevalence data. Green curve and ribbon indicate the median and 95% CrIs estimated by the ensemble model. Dots and lines represent the mean and 95% binomial confidence interval reported by the published seroprevalence data. For countries with > 1 seroprevalence surveys, black dots and lines correspond to the study-1 as referenced in Figure 2 and Supplementary Table S1, whereas pink dots and lines correspond to the study-2 (e.g. Belgium-2). Blue shaded regions indicate the start and end dates of sampling for each seroprevalence survey.

## Heterogeneities in ≥65 mortality

Using our model framework we estimate the number of deaths expected in the absence of nursing home transmission in those aged ≥65 years, given the reported number of deaths in younger age groups. These estimates can be compared to the reported number of COVID-19 deaths in ≥65 year olds (Figure 4A). We find that many countries in South America had significantly fewer reported deaths in individuals ≥65 years than expected, consistent with under-reporting of COVID-19 deaths amongst elderly individuals. For example, we find that in Ecuador there are 231 fewer reported deaths per 100,000 in those ≥65 years than expected (95%CrI: 211-253), equivalent to approximately 3,000 missing deaths. While lower infection attack rates in elderly populations due to reduced contacts and/or successful shielding policies may also explain lower mortality amongst older individuals, in sensitivity analyses we show that for some countries unrealistically low infection attack rates amongst ≥65 year olds compared to the rest of the population would be required to reconcile the reported number of deaths in these age-groups (Supplementary Figure S3).

**Figure 4.**
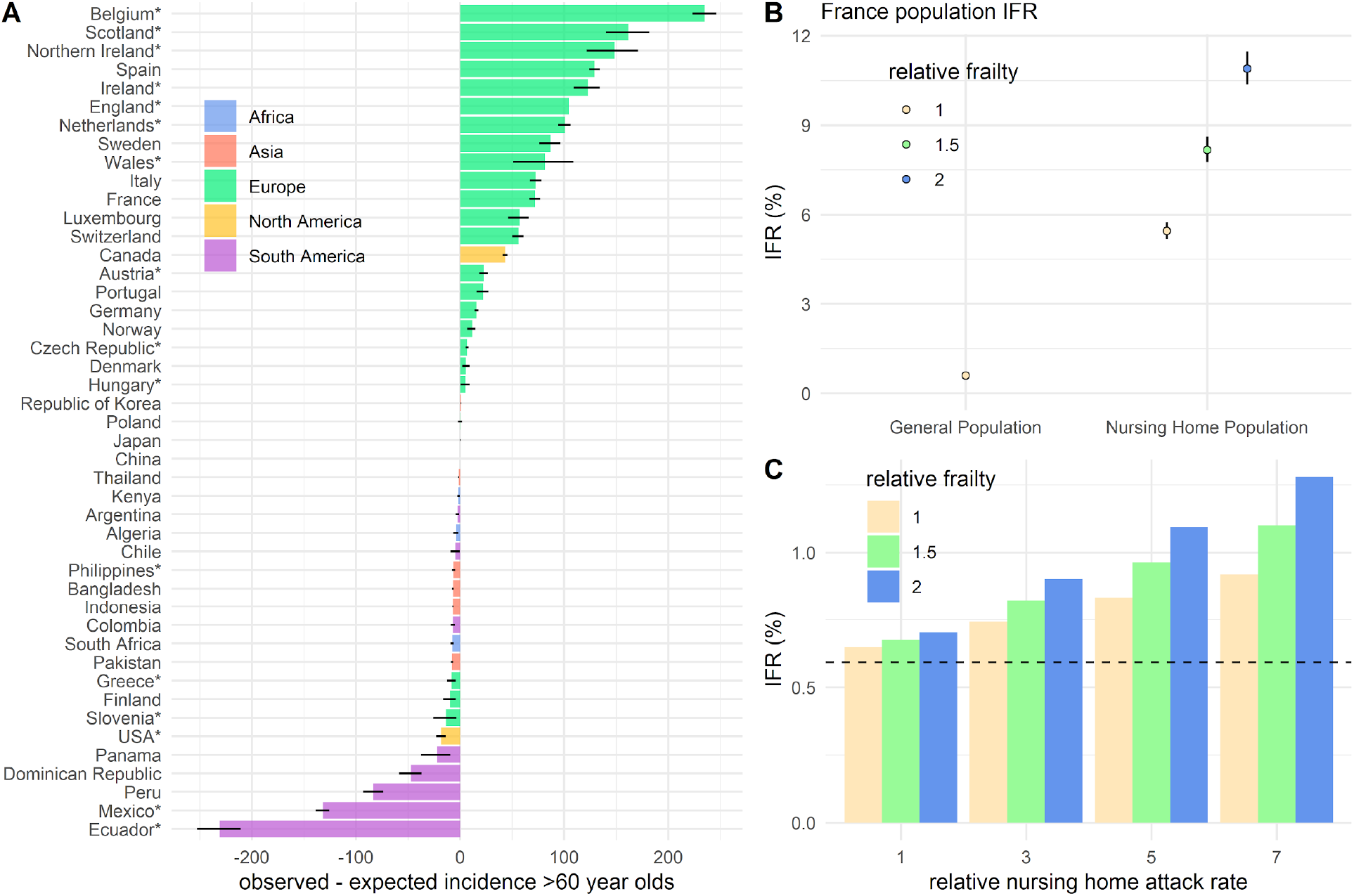
Infection fatality patterns amongst ≥60s. (A) Difference between the reported and expected incidence of COVID-19 deaths per 100,000 population amongst ≥60s or ≥65s in each country. Coloured bars represent the median difference and black lines represent 95% credible intervals (CrIs). Countries labelled with an asterisk * indicate where the number of deaths were reconstructed for ages 65+, to align with the reported age-groups for each country. (B) Population-weighted IFRs for the general population and nursing home residents, using France as a reference population. The relative frailty of nursing home residents is assumed to be 1 (yellow), 1.5 (green), or 2 (blue). Dots and lines indicate the median and 95% CrIs estimated by the ensemble model. (C) Population-weighted IFR in France, estimated with different assumed infection attack rates and relative frailty for nursing home residents relative to the general population. The black dashed line represents the median population-weighted IFR estimated when assuming a zero infection attack rate amongst nursing home residents.

By contrast, for many European countries we observe a higher incidence of deaths in older individuals than expected (Figure 4A). This is consistent with the large proportion of reported COVID-19 deaths attributable to outbreaks in nursing homes, highlighting the enormous burden experienced by these communities in many higher-income countries. Using France as a reference population, we use the age and sex distribution of nursing home residents to derive a population-weighted IFR of 5.45% (95%CrI: 5.18-5.74%) among French nursing home residents, assuming equal frailty of individuals in nursing homes and the general population of the same age and gender (Figure 4B). Using this estimate of the IFR would suggest that 29.05% of the nursing home population had been infected by 30th May 2020 (95%CrI: 27.60-30.58%), a 6.14 fold higher infection attack rate than the general population (Supplementary Methods S3). Assuming individuals in nursing homes are twice as frail as the general population would imply a relative infection attack rate of 3.07 or 14.52% (95%CrI: 13.80-15.29%) of the nursing home population infected. In our baseline model we have derived IFR estimates assuming the absence of excess nursing home transmission and mortality so as to facilitate robust comparisons of IFR and general population transmission across settings. However, we demonstrate that where high rates of infection have occurred amongst nursing home residents, population IFR estimates will be significantly greater than in scenarios where these populations have been successfully shielded or experienced little exposure (Figure 4C). For example, in France, including deaths in nursing homes, increases the IFR from 0.60% for the general population (95%CrI: 0.57-0.63%) to 0.88% overall (95%CrI: 0.84-0.93%), assuming equal frailty. This highlights the complexity in comparing headline IFR estimates across populations where very different levels of transmission may have occurred in these hyper-vulnerable communities.

## Discussion

Seroprevalence surveys have, to date, shown inconsistent patterns in age-specific attack rates (Figure S7). Contact patterns are likely to have changed significantly over the course of the pandemic, particularly for older individuals who may have further reduced social contacts as part of shielding interventions or natural behavioural change. To attempt to account for this, in our baseline model, we have assumed equal infection attack rates amongst <65s and a relative attack rate of 0.7 amongst individuals aged ≥65. Sensitivity analyses where we assume constant attack rates across ages provides similar estimates (Figure S6). Here we have used national reporting systems of COVID-19 associated deaths, however, other approaches exist. For example, excess deaths have been used to estimate SARS-CoV-2 burden, though these are rarely available by age and sex. We find a consistent relationship between the total number of reported COVID-19 deaths and excess deaths for 21 countries, where both are available, with the notable exceptions of Peru and Ecuador, consistent with our finding that these two countries have fewer reported deaths than expected (Figure S1).

Translating the number of COVID-19 deaths into estimates of the number of infections requires careful consideration of fatalities that may have occurred from outbreak events in highly vulnerable populations. This study shows the valuable information provided by the age distribution of COVID-19 deaths and how deaths in those aged <65 in particular can be used to provide simple, robust estimates of the underlying proportion of the population that have been infected. This is of critical use in a context where most infections are unobserved. Our approach allows us to identify countries where excess transmission in nursing home populations is likely to have occurred, far exceeding that of the general population, and locations where deaths in the elderly population are likely to be under-reported. The results and modelling framework we present demonstrate how age-specific death data alone can be used to reconstruct the underlying level of infection. This approach could be applied at sub-national scale and may be of particular use in settings where there do not exist the resources to carry out large, representative seroprevalence studies.

## Data Availability

All code and data necessary to reproduce this analysis are available at 491 https://github.com/meganodris/International-COVID-IFR.

## Methods

### Data

#### Age- and sex-specific COVID-19 fatality data

We collated national-level age-stratified COVID-19 death counts from official government and department of health webpages and reports for 45 countries. Where available, the stratification by both age and sex were used. Sub-national age-stratified death counts were additionally collated for regions where seroprevalence surveys had been conducted. For countries/regions where information on age was missing for a subset of deaths, we assumed the age-distribution of the missing subset to be the same as that of the deaths with available age data. Information on age was missing for 28% of deaths in Belgium and 29% in Spain. In addition, the time series of daily reported deaths from each country/region were obtained from the COVID-19 Data Repository by the Center for Systems Science and Engineering (CSSE) at Johns Hopkins University^16^.

#### Seroprevalence studies

We used data from 18 SARS-CoV-2 seroprevalence surveys from 15 countries/regions where the results were representative of the general population and where age-stratified death data were also available, shown in Figure 1A and Supplementary Table S1. In the ensemble model we consider only the 15 national-level seroprevalence surveys, representing 12 countries. Where reported, estimates of seroprevalence adjusted for assay performance and/or population demographics were used preferentially to unadjusted estimates (Supplementary Table S1).

### Model

We combined age- and sex-specific COVID-19 death data from 45 countries with data from 15 seroprevalence surveys, to jointly infer the age- and sex-specific IFRs and country-specific cumulative probabilities of infection. Age- and sex-specific IFRs were estimated in 5-year age-groups, with individuals aged 80+ considered as a single age group. Let *N_c,a,s_* be the population size for the age group *a* of sex *s* in country *c*. The expected number of deaths for the age group *a* of sex *s* in country *c*, *D_c,a,s_* is estimated as shown in equation 1, which we assume to follow a Poisson distribution. *Λ_c_* denotes the cumulative probability of infection in country *c*, *δ_a_* the relative probability of infection in age-group *a*, and *IFR_a,s_*the infection fatality ratio of age-group *a* and sex *s*.

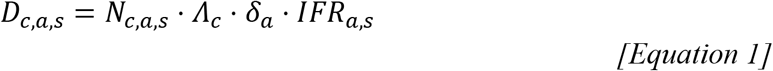

The expected number of deaths estimated by 5-year age-groups were summed to match the corresponding age-groups of observed deaths when reported in coarser age-groups. We fit exclusively to the reported number of deaths for age groups <65 years for each country (i.e. including all age-groups where the upper bound is <65 years). IFRs for age groups ≥65 were derived from age-specific death data reported by the Office for National Statistics (ONS) in England^17^, which allows us to exclude deaths among nursing home residents (Supplementary Methods S2). As an external validation, we apply these IFRs to reported death data for a subset of 13 countries where an adjustment for deaths occurring in nursing homes could be applied (Supplementary Methods S2).

To align estimates of the cumulative probability of infection, *Λ_c_*, with data from seroprevalence surveys, we used daily time-series of reported deaths to infer the timing of infections and subsequent seroconversions. We assumed a gamma distributed delay between onset and death with mean of 17.8 and standard deviation of 8 days^2^ and a gamma distributed delay between onset and seroconversion with a mean of 10 and standard deviation of 8 days^18^. We derive the approximated seroprevalence at a given survey period *t*, *λ_c,t_*, as shown in equation 2, where *S_c,i_* is the inferred number of seroconversions in country *c* on day *i*, *D_c,i_* the number of new deaths reported in country *c* on day *i*, and *T_c_* is the date of reporting of the age-stratified cumulative death data.

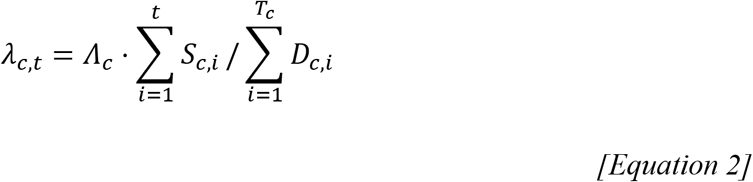

For each seroprevalence survey the expected number of seropositive individuals in country *c* at sampling period *t*, *NPos_c,t_*, is assumed to follow a Binomial distribution as shown in equation 3, where *NSamples_c,t_* is the number of serological samples taken in country *c* at time *t*^19^.

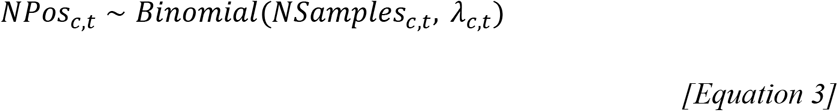

Where reported, seroprevalence estimates adjusted for test performance and/or population demographics were used preferentially to unadjusted values (Supplementary Table S1). To investigate the contribution of different serological studies to the likelihood the model was fit separately to each individual serostudy, including an additional 3 subnational seroprevalence studies (Supplementary Table S1). All parameters were estimated in a Bayesian framework using RStan^20^.

### Code and Data Availability

All code and data necessary to reproduce this analysis are available at https://github.com/meganodris/International-COVID-IFR.

## Supplementary Information

**Table S1.** Seroprevalence studies

| Country/Region | Study Period | N | N Positive (%) | Population | Age | Source |
| --- | --- | --- | --- | --- | --- | --- |
| Belgium | 03/03/2020 – 05/04/2020 | 3,910 | 113 (2.9%)<sup>a</sup> | General | 0-101 | (1) |
|  | 20/04/2020 – 26/04/2020 | 3,397 | 204 (6.0%)<sup>a</sup> | General | 0-101 | (1) |
|  | 14/04/2020 – 16/04/2020 | 900 | 43 (4.8%) | Blood donors | 18-75 | (2) |
|  | 27/04/2020 – 29/04/2020 | 900 | 42 (4.7%) | Blood donors | 18-75 | (2) |
|  | 11/05/2020 – 13/05/2020 | 900 | 42 (4.7%) | Blood donors | 18-75 | (2) |
| Czech Republic | 23/04/2020 – 01/05/2020 | 25,549 | 107 (0.4%) | General | 18-89 | (3) |
| Denmark | 06/04/2020 – 08/04/2020 | 4,072 | 61 (1.5%)<sup>b</sup> | Blood donors | 17-69 | (4) |
|  | 14/04/2020 – 19/04/2020 | 5,326 | 101 (1.9%) | Blood donors | 17-69 | (5) |
|  | 20/04/2020 – 26/04/2020 | 5,820 | 93 (1.6%) | Blood donors | 17-69 | (5) |
|  | 27/04/2020 – 03/05/2020 | 5,422 | 130 (2.4%) | Blood donors | 17-69 | (5) |
| England | 26/04/2020 – 13/06/2020 | 1,757 | 95 (5.4%) | General | 16+ | (6) |
| Finland | 13/04/2020 – 19/04/2020 | 362 | 9 (2.5%) | General | 18-69 | (7) |
|  | 20/04/2020 – 26/04/2020 | 674 | 17 (2.5%) | General | 18-69 | (7) |
|  | 27/04/2020 – 03/05/2020 | 426 | 12 (2.8%) | General | 18-69 | (7) |
|  | 04/05/2020 – 10/05/2020 | 514 | 8 (1.6%) | General | 18-69 | (7) |
|  | 11/05/2020 – 17/05/2020 | 401 | 4 (1.0%) | General | 18-69 | (7) |
|  | 18/05/2020 – 24/05/2020 | 210 | 9 (4.3%) | General | 18-69 | (7) |
|  | 25/05/2020 – 31/05/2020 | 178 | 5 (2.8%) | General | 18-69 | (7) |
|  | 01/06/2020 – 07/06/2020 | 190 | 6 (3.2%) | General | 18-69 | (7) |
| France | 09/03/2020 – 15/03/2020 | 3,084 | 36 (1.1%) | General | 0+ | (8) |
|  | 06/04/2020 – 12/04/2020 | 3,221 | 208 (6.7%) | General | 0+ | (8) |
| Geneva | 06/04/2020 – 10/04/2020 | 341 | 16 (4.8%)<sup>a,b</sup> | General | 5+ | (9) |
|  | 14/04/2020 – 17/04/2020 | 469 | 40 (8.5%)<sup>a,b</sup> | General | 5+ | (9) |
|  | 20/04/2020 – 24/04/2020 | 577 | 63 (10.9%)<sup>a,b</sup> | General | 5+ | (9) |
|  | 27/04/2020 – 02/05/2020 | 604 | 40 (6.6%)<sup>a,b</sup> | General | 5+ | (9) |
|  | 04/05/2020 – 09/05/2020 | 775 | 84 (10.8%)<sup>a,b</sup> | General | 5+ | (9) |
| Luxembourg | 15/04/2020 – 05/05/2020 | 1,862 | 35 (1.9%) | General | 18-79 | (10) |
| Netherlands | 01/04/2020 – 15/04/2020 | 7,361 | 200 (2.7%) | Blood donors | 18-72 | (11) |
|  | 01/04/2020 – 17/04/2020 | 2,096 | 75 (3.6%) | General | 2+ | (12) |
| New York City | 19/04/2020 – 28/04/2020 | 5,946 | 1,350 (14.0%)<sup>b</sup> | General | 18+ | (13) |
| New York State | 19/04/2020 – 28/04/2020 | 15,101 | 2,114 (22.7%)<sup>b</sup> | General | 18+ | (13) |
| Scotland | 17/03/2020 – 17/03/2020 | 500 | 0 (0.0%) | Blood donors | 18-75 | (14) |
|  | 21/03/2020 – 23/03/2020 | 500 | 6 (1.2%) | Blood donors | 18-75 | (14) |
| Spain | 17/04/2020 – 11/05/2020 | 61,075 | 3,054 (5.0%) | General | 0+ | (15) |
|  | 18/05/2020 – 01/06/2020 | 63,564 | 3,305 (5.2%) | General | 0+ | (15) |
| Slovenia | 10/04/2020 – 10/04/2020 | 1,318 | 41 (3.1%) | General | 0-99 | (16, 17) |
| Sweden | 27/04/2020 – 03/05/2020 | 1,200 | 66 (5.5%)<sup>a,c</sup> | Hospital outpatients | 0-95 | (18) |
|  | 04/05/2020 – 10/05/2020 | 1,200 | 58 (4.8%)<sup>a,c</sup> | Hospital outpatients | 0-95 | (18) |
|  | 11/05/2020 – 17/05/2020 | 1,200 | 73 (6.1%)<sup>a,c</sup> | Hospital outpatients | 0-95 | (18) |
|  | 18/05/2020 – 24/05/2020 | 1,200 | 76 (6.3%)<sup>a,c</sup> | Hospital outpatients | 0-95 | (18) |

<sup>a</sup> Adjusted for population demographics.
<sup>b</sup> Adjusted for assay performance.
<sup>c</sup> The number of samples is reported as approximately 1,200 samples per week<sup>18</sup>. The absolute number of positive samples was inferred using the reported seroprevalence values assuming sample sizes of exactly 1,200 per week.

## Supplementary Methods

### S1. Relative risk of COVID-19 death by age

To explore the risk of reported COVID-19 death by age in each country age-specific relative risks (RR) of death were calculated as shown in equation 1, where *D_c,a_* and *N_c,a_* are the country and age-specific number of deaths and population size, respectively. The age-group 55-59 was chosen as the preferential reference group as it is less likely to be influenced by deaths associated with outbreaks in nursing home settings. As the reported age-groups varied by country, the age group with an upper bound of 59 was chosen as the reference group where possible. Where this was not an available age-group, the age-group with an upper bound of 64 was selected as the reference.

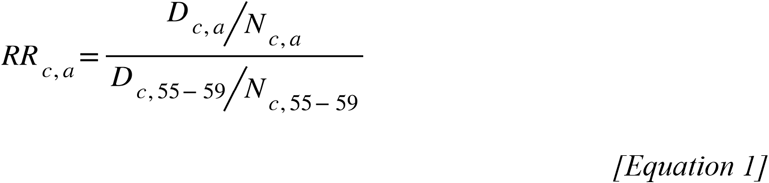

### S2. Inferring IFR estimates amongst >65s

We fit our model exclusively to deaths <65 years. We use age-specific death data for England, reported by the Office of National Statistics (ONS)^19^, to derive IFR estimates for age groups ≥65, adjusted for nursing home deaths and assuming a baseline relative infection attack rate of 0.7 for those aged 65+ relative to individuals aged <65 as shown in Equation 2.

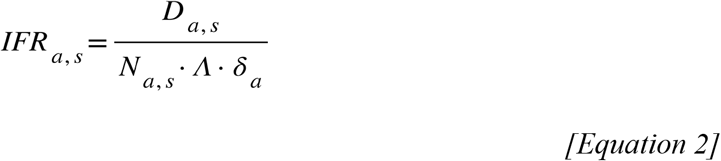

Here, *D_a,s_* is the number of age and sex-specific non-nursing home COVID-19 deaths, *N_a,s_* is the age and sex-specific population size, *Λ* is the cumulative probability of infection and *δ_a_* is the age-specific relative infection attack rate. The age- and sex-specific number of non-nursing home COVID-19 deaths were calculated by assuming that all COVID-19 deaths that occurred in nursing homes were aged 65+ and that the age-sex-distribution of these deaths follows the same age-sex-distribution as all COVID-19 deaths ≥65 years. We applied this adjustment to an additional 13 countries where the proportion of COVID-19 deaths attributable to nursing homes had been reported, assuming the cumulative proportions to be constant in time (Table S2). In the case of France, deaths that occurred in nursing homes are reported separately to those that occurred in hospital. As minimal proportions of reported hospitalised deaths are expected to be attributable to nursing home residents, we treat the reported hospitalised deaths in France as non-nursing home deaths. To assess the generalizability of IFRs ≥65 derived from this data, we apply them to the 13 additional countries and find that they can reconstruct the number of non-nursing home deaths relatively well in these countries (Figure 1C).

**Table S2.** The proportion of reported COVID-19 deaths attributable to nursing home/long-term care (LTC) settings for 13 countries.

| Country | Date of Reporting | % (LTC/Total COVID-19 deaths) |
| --- | --- | --- |
| Denmark | 24/04/2020 | 33.76% (133/394) |
| England | 01/05/2020 | 26.06% (8,280/31,777) |
| Finland | 28/05/2020 | 45.05% (141/313) |
| Germany | 03/05/2020 | 36.11% (2,401/6,649) |
| Hungary | 18/04/2020 | 19.19% (33/172) |
| Ireland | 06/05/2020 | 62.33% (857/1,375) |
| Northern Ireland | 08/05/2020 | 45.74% (274/599) |
| Norway | 02/05/2020 | 60.19% (127/211) |
| Portugal | 23/04/2020 | 39.88% (327/820) |
| Scotland | 10/05/2020 | 44.76% (1,438/3,213) |
| Sweden | 14/05/2020 | 56.47% (1,637/2,899) |
| Switzerland | 12/05/2020 | 53.00% (927/1,749) |
| Wales | 01/05/2020 | 26.37% (413/1,566) |

### S3. Population IFR estimates and nursing home transmission

To account for nursing home deaths in the estimation of IFR, we define 2 distinct populations - that of nursing home residents, *N_NH_*, and that of the general population excluding nursing home residents, *N*_g_. For each of these populations we derive single population-weighted IFR values, *IFRg* and *IFR_NH_* (Figure 4B), using the age and sex-specific IFR estimates produced by the ensemble model and the demographic distributions of each population as shown in equations 3 and 4. *IFR_a,s_* is the age and sex-specific IFR estimates from the ensemble model, *α_a,s_* is the proportion of the population in age group *a* and sex *s*, and *γ* is the frailty of nursing home residents relative to that of the general population of the same age and sex.

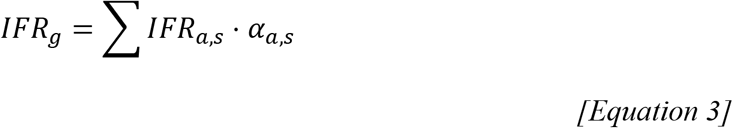

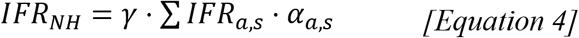

To demonstrate how varying levels of transmission in nursing home settings can affect estimates of IFR (Figure 4C) we apply a simplified calculation of the overall IFR, shown in equation 5. Here, the total number of COVID-19 deaths in nursing home settings, *D_NH_*, is varied through values of the nursing home infection attack rate, *λ_NH_*, and relative frailty, *γ*, as shown in equation 6, where values of *γ*>1 represents increased frailty of nursing home residents relative to that of the general population. The total number of COVID-19 deaths in the general population, *D*_g_, the infection attack rate of the general population, *λ*_g_, and the population sizes of both the general and nursing home populations, *N_g_* and *N_NH_* remain fixed.

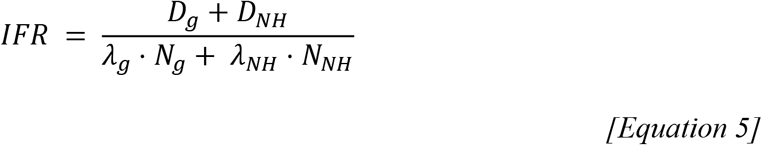

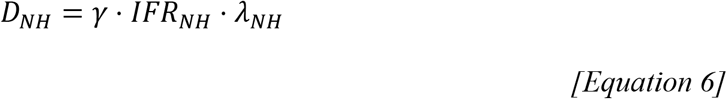

Using the total number of reported COVID-19 deaths attributed to nursing home residents the infection attack rate in nursing home settings can be approximated, shown in Equation 7, under different scenarios of the relative frailty of nursing home residents, *γ*.

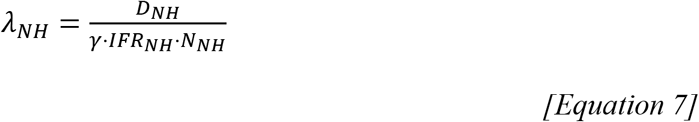

## Supplementary Discussion

### S4. Excess Deaths

In our main analysis we use the number of COVID-19 deaths reported by individual countries. As the completeness of reporting of these data across countries is unclear, we investigate the number of excess deaths as compared to previous years, where available, for a subset of 21 countries. We use data from The Economist “covid-19-excess-deaths-tracker” repository which collates data from statistical bureaus, health ministries and government departments20. Of the 21 countries for which excess death data were available we find largely consistent numbers of excess deaths and reported COVID-19 deaths used in our analysis, per 100,000 population (Figure S1). Two outliers include Ecuador and Peru, where the number of excess deaths is much larger than the reported number of COVID-19 deaths, consistent with where our model predicts the largest difference in expected and reported COVID-19 deaths amongst ≥65s (Figure 4).

**Figure S1.**
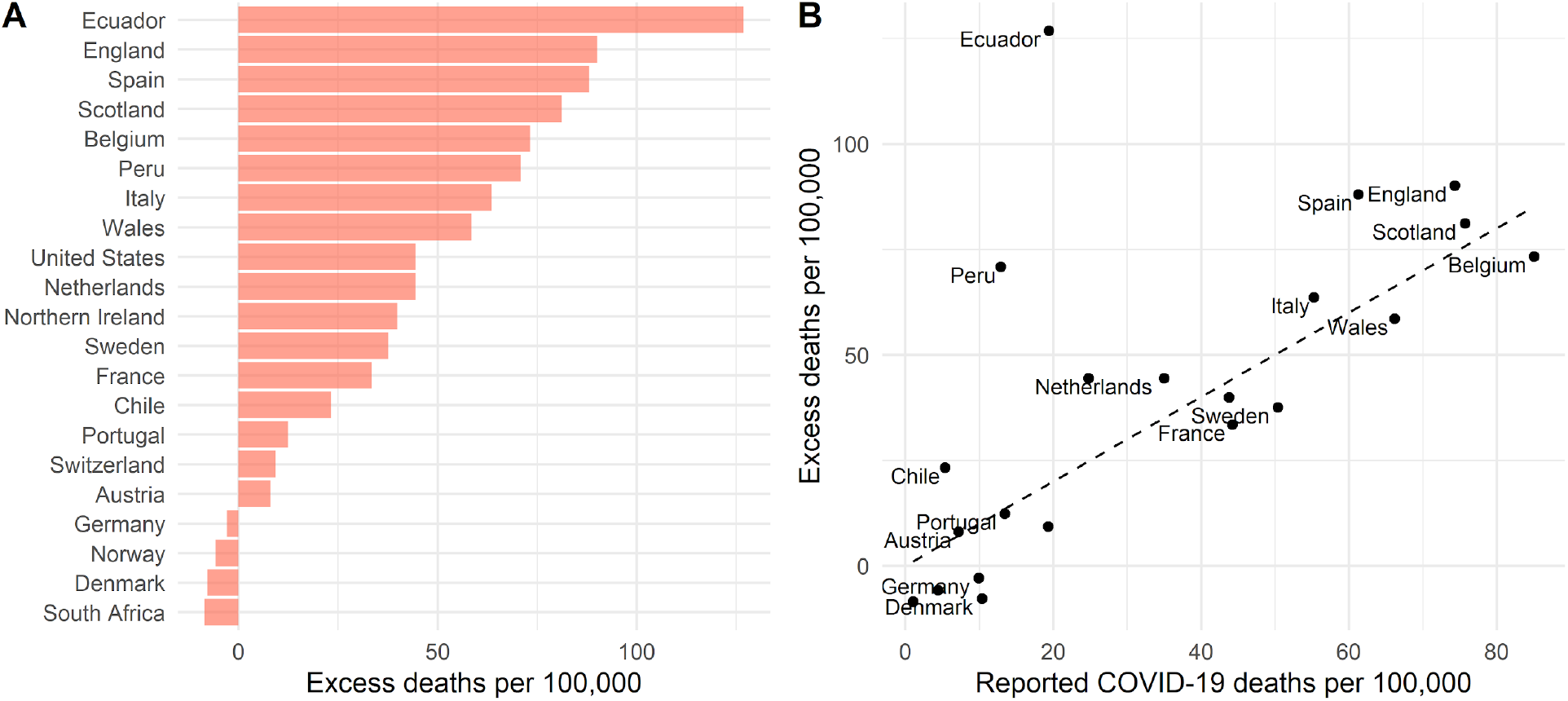
(A) Excess deaths per 100,000 population for 21 countries, January-May 2020, as compared to averages for the same time period of previous years. (B) Relationship between excess deaths and reported COVID-19 deaths per 100,000 population. The dashed line represents the x=y line, highlighting where excess deaths would equal reported COVID-19 deaths per 100,000.

**Supplementary Figure S2.**
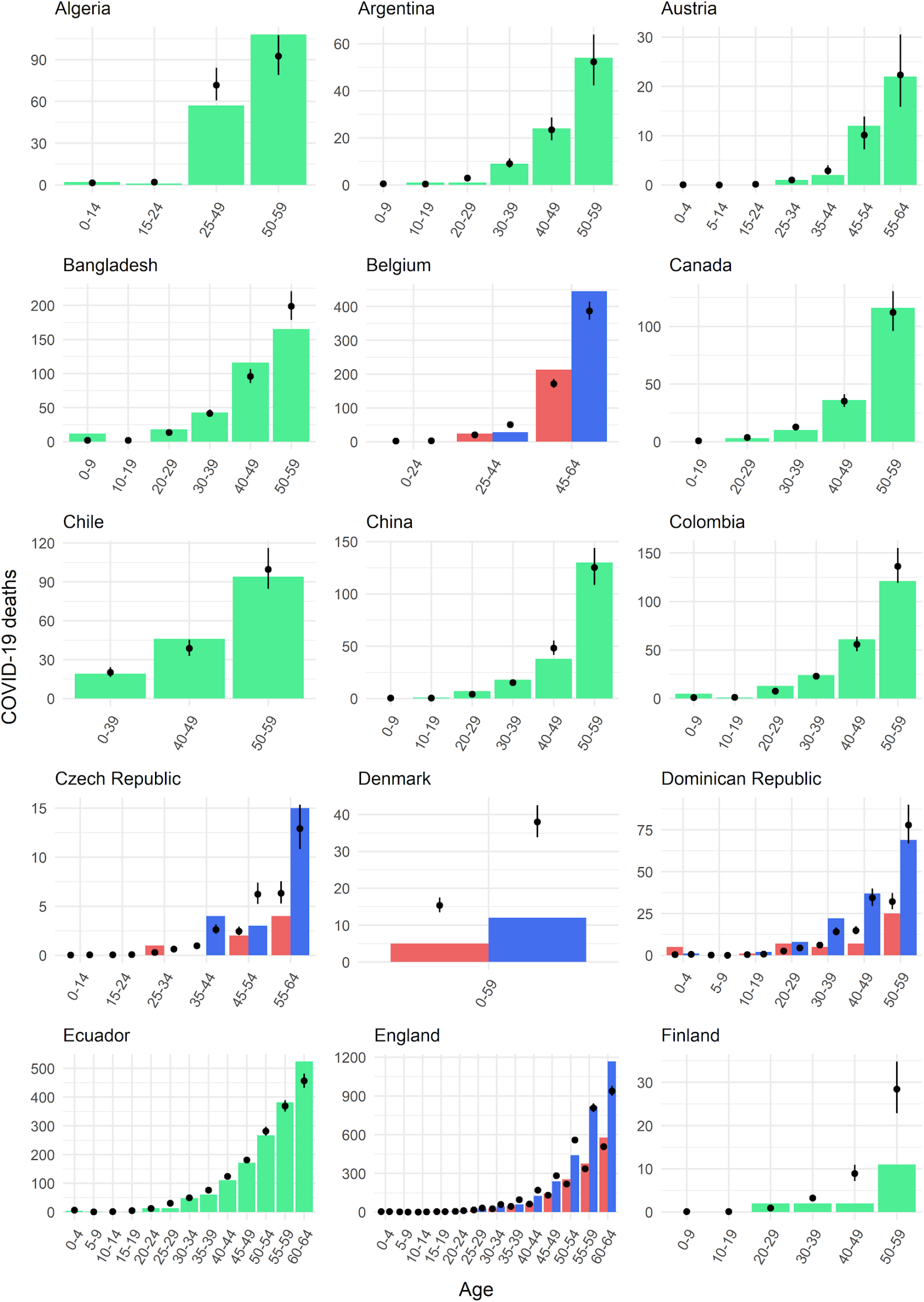
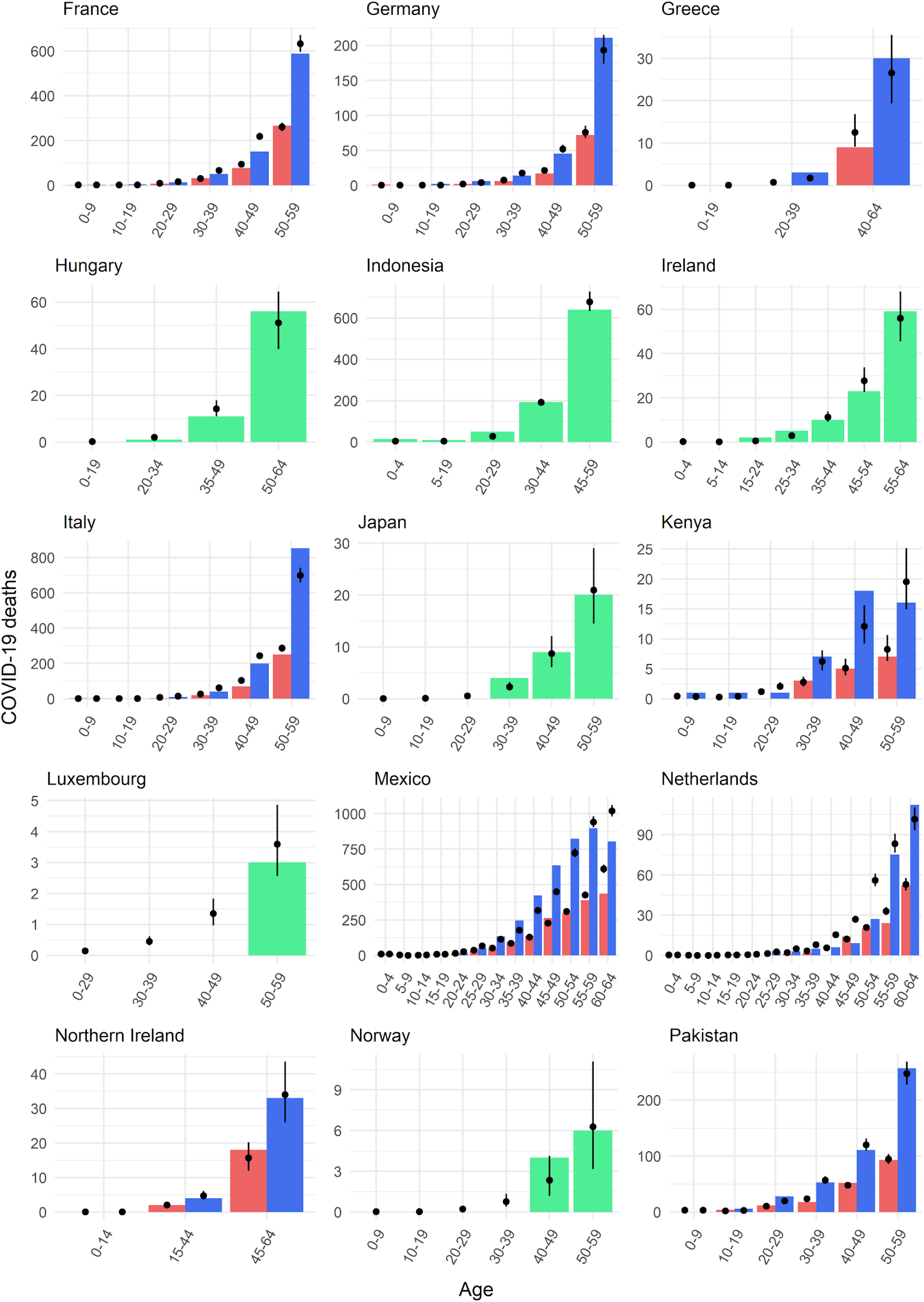
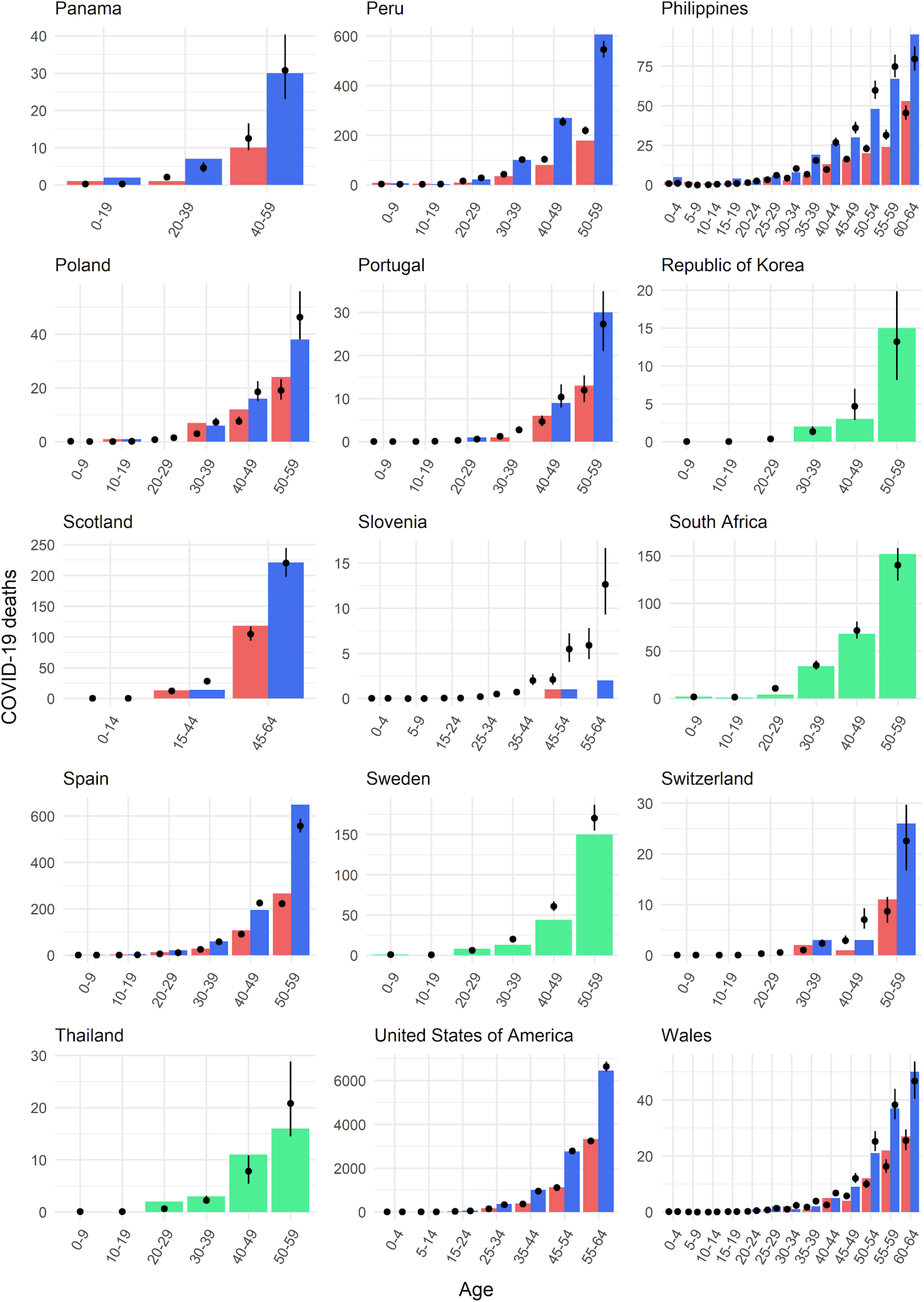
Fit to age-specific death data in age groups <65. Coloured bars represent the observed age-specific number of deaths in each country (blue=male, red=female, green=both). Black points and lines represent the median and 95% credible interval model estimates.

**Supplementary Figure S3.**
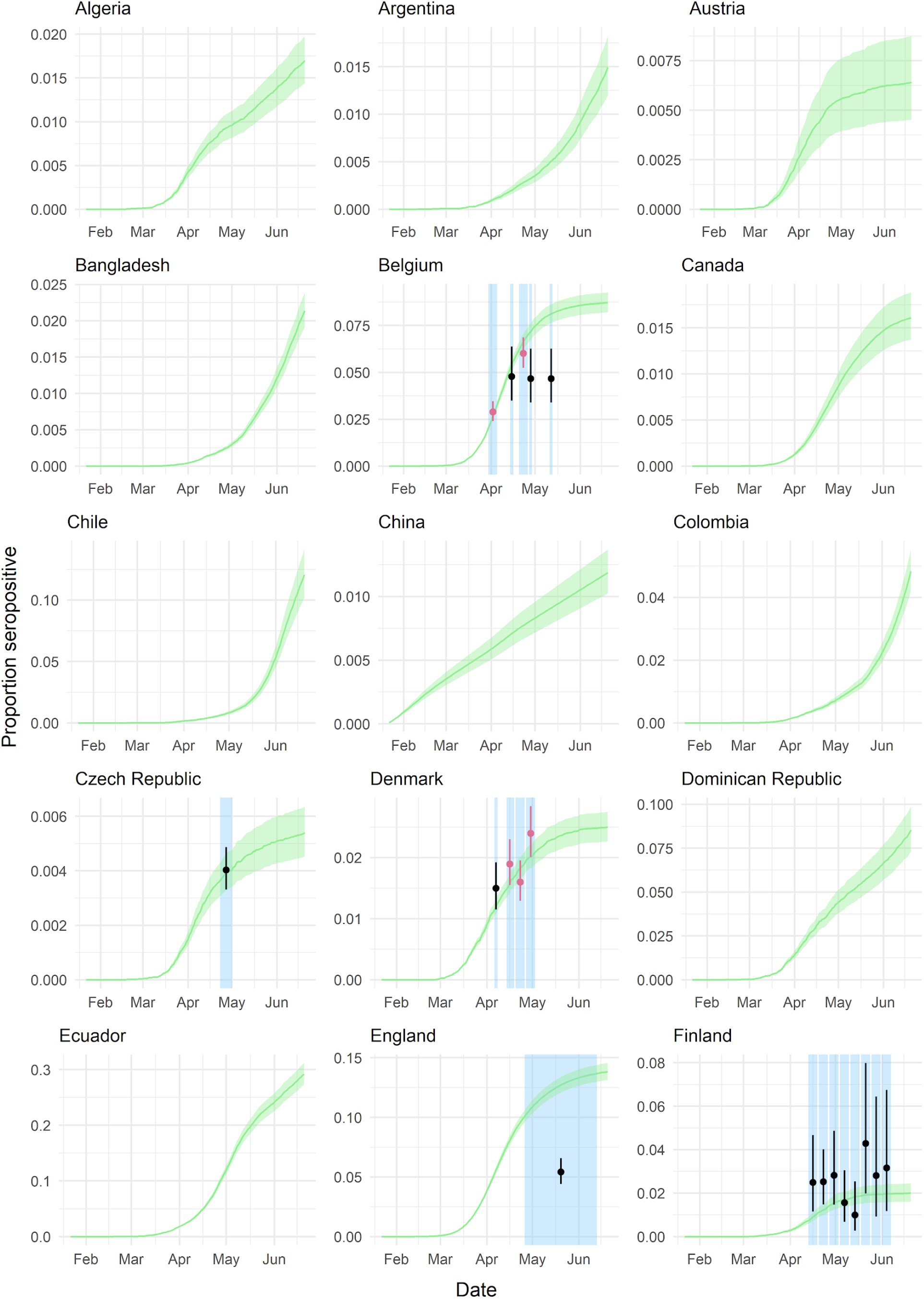
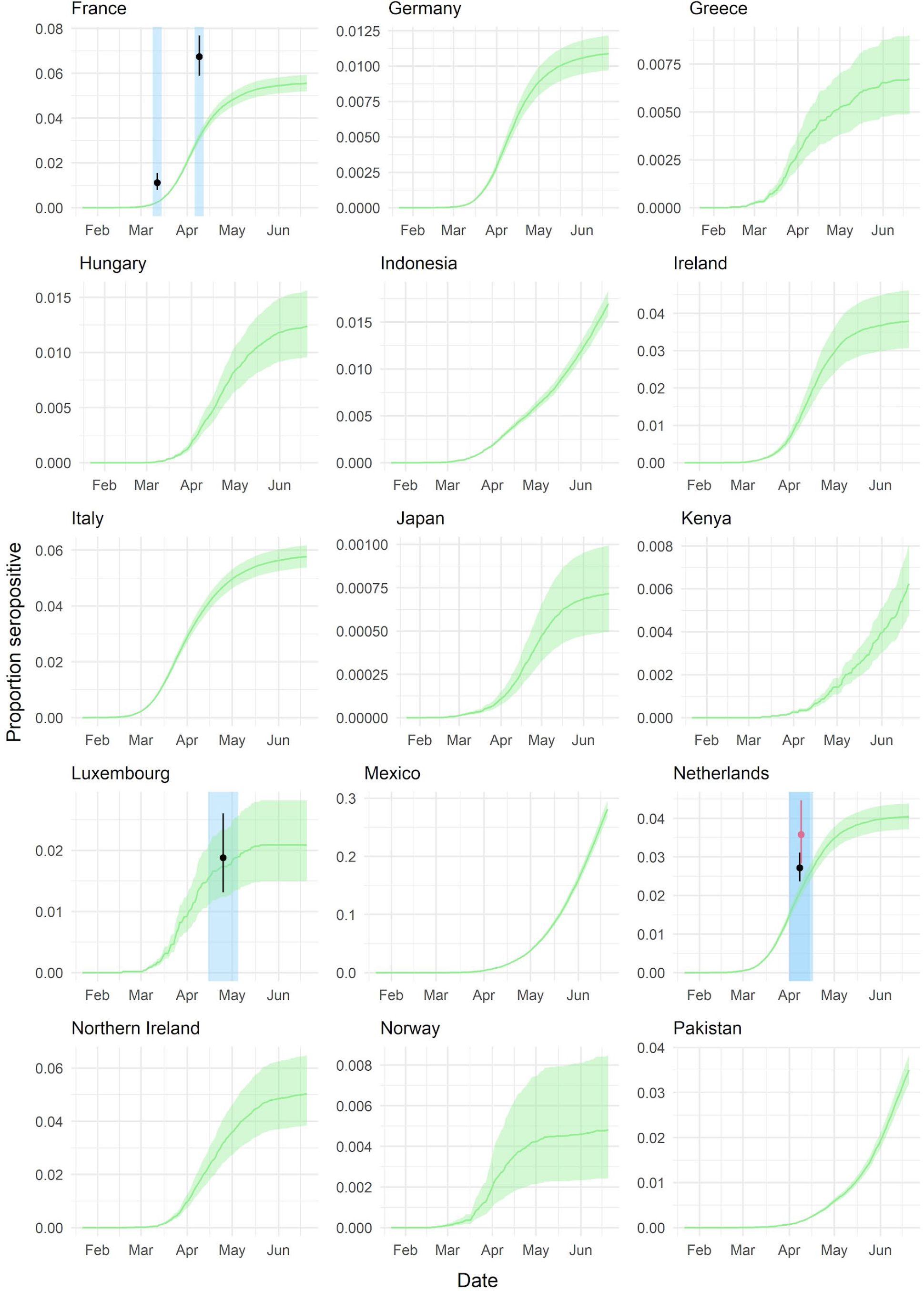
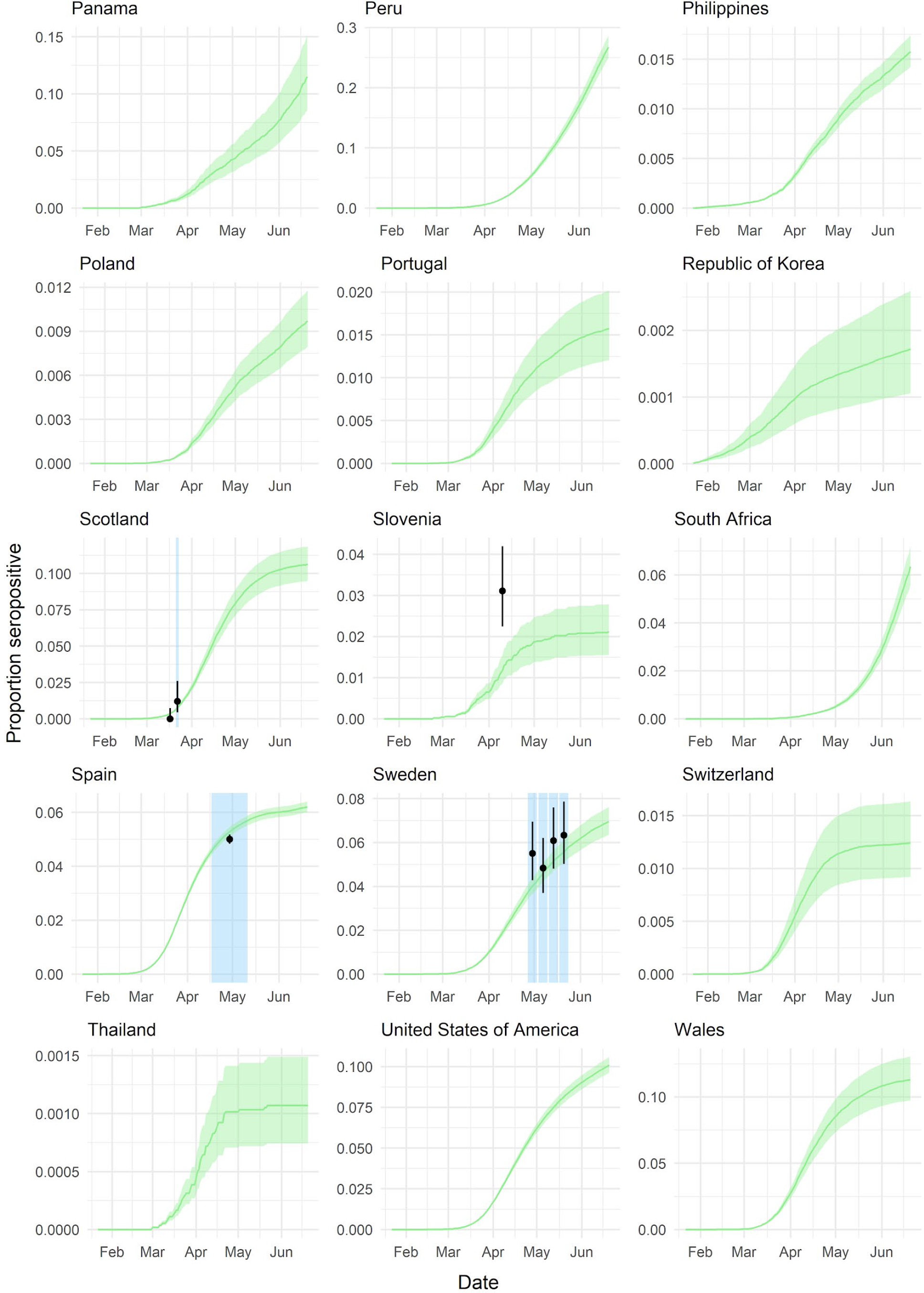
Estimates of the proportion seropositive over time. The green line and ribbons represent the median and 95% credible interval estimates of the proportion seropositive over time. Coloured points and lines represent the proportion seropositive as reported by seroprevalence studies and the blue shading shows the timing of each seroprevalence sampling period.

**Figure S4.**
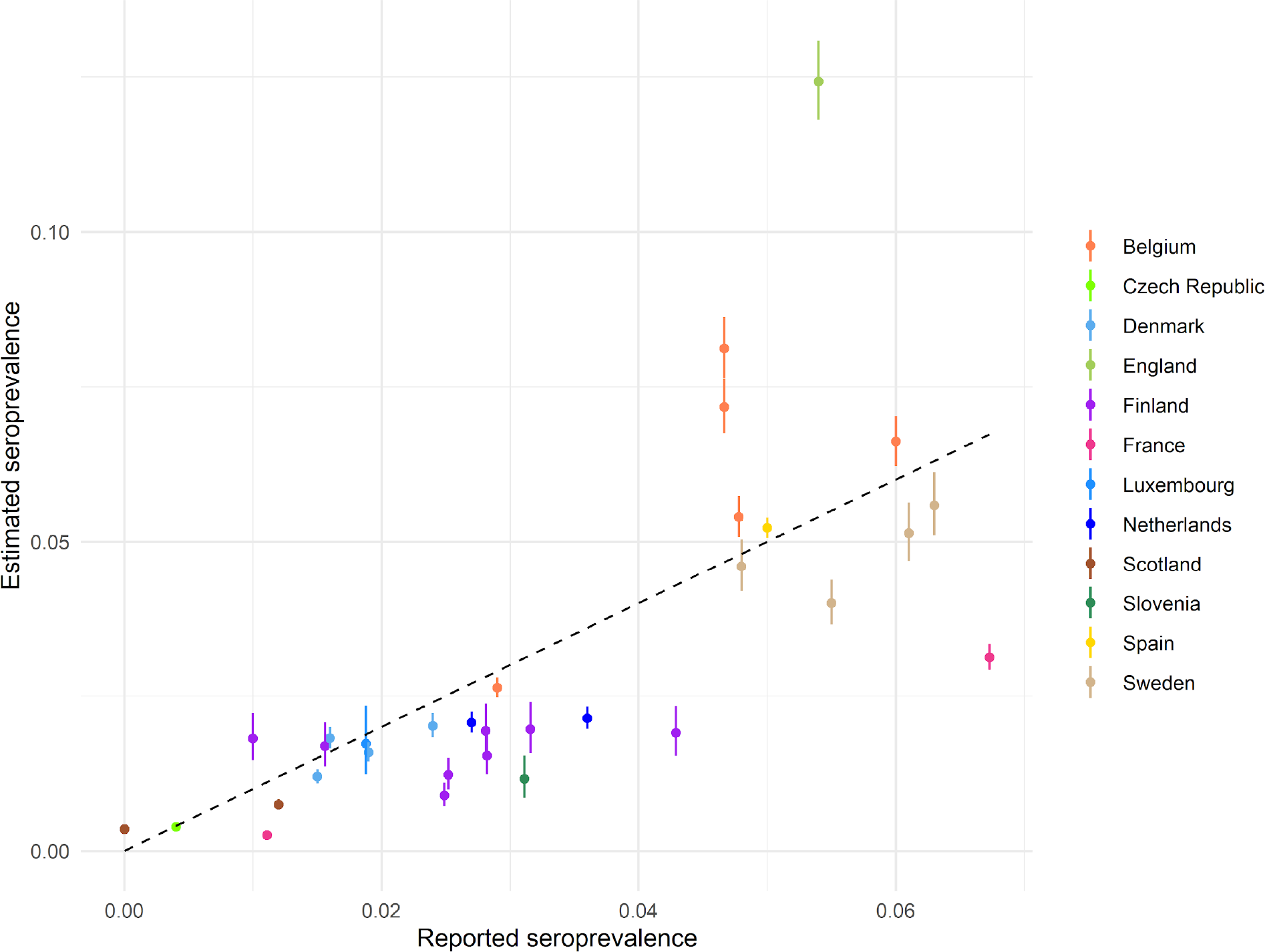
Ensemble model fit to reported seroprevalence values. Dots and lines represent median and 95% credible interval estimates from the ensemble model. The black dashed line indicates the x=y line, highlighting where reported and estimated seroprevalence would be equal.

**Figure S5.**
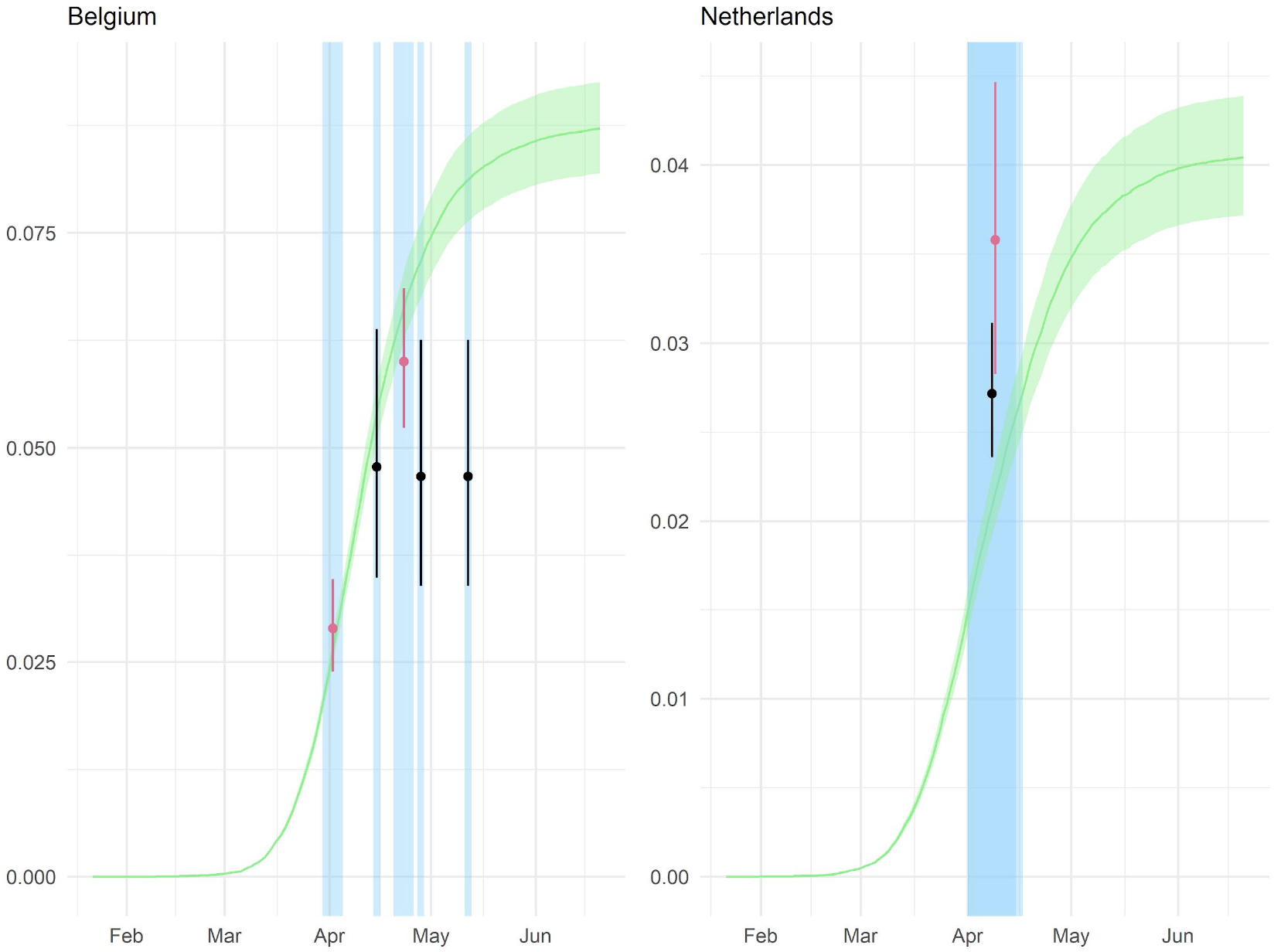
Estimated and reported seroprevalence over time where both blood donor and general population seroprevalence surveys were conducted. Green line and ribbon indicate the median and 95% credible interval estimates of the ensemble model. Blue shading represents the timing of sampling of each reported seroprevalence estimate. Dots and lines show the mean and 95% binomial confidence intervals of the reported seroprevalence, where black represents studies conducted amongst the general population and pink represents studies conducted in blood donor samples.

**Table S4.** Ensemble model age- and sex-specific infection fatality ratio estimates. Median and 95% credible intervals (CrI), for males, females and the mean of male and female estimates.

| Age | Male<br>Median (95%CrI) % | Female<br>Median (95%CrI) % | Mean<br>Median (95%CrI) % |
| --- | --- | --- | --- |
| 0-4 | 0.002 (0.001-0.002) | 0.002 (0.001-0.002) | 0.002 (0.001-0.002) |
| 5-9 | 0.000 (0.000-0.000) | 0.001 (0.000-0.000) | 0.000 (0.000-0.000) |
| 10-14 | 0.001 (0.000-0.001) | 0.000 (0.001-0.002) | 0.000 (0.000-0.001) |
| 15-19 | 0.002 (0.001-0.002) | 0.001 (0.001-0.002) | 0.002 (0.001-0.002) |
| 20-24 | 0.005 (0.004-0.006) | 0.003 (0.002-0.004) | 0.004 (0.003-0.004) |
| 25-29 | 0.012 (0.010-0.014) | 0.007 (0.005-0.008) | 0.009 (0.008-0.011) |
| 30-34 | 0.024 (0.021-0.026) | 0.010 (0.009-0.012) | 0.017 (0.015-0.019) |
| 35-39 | 0.040 (0.037-0.044) | 0.018 (0.016-0.020) | 0.029 (0.027-0.031) |
| 40-44 | 0.077 (0.072-0.083) | 0.028 (0.025-0.031) | 0.053 (0.049-0.056) |
| 45-49 | 0.118 (0.110-0.126) | 0.053 (0.049-0.058) | 0.086 (0.081-0.091) |
| 50-54 | 0.223 (0.210-0.236) | 0.084 (0.078-0.091) | 0.154 (0.0146-0.162) |
| 55-59 | 0.344 (0.326-0.362) | 0.138 (0.130-0.148) | 0.241 (0.229-0.253) |
| 60-64 | 0.473 (0.446-0.500) | 0.246 (0.213-0.262) | 0.359 (0.340-0.379) |
| 65-69 | 0.868 (0.824-0.913) | 0.417 (0.396-0.439) | 0.642 (0.610-0.676) |
| 70-74 | 1.445 (1.373-1.521) | 0.707 (0.671-0.744) | 1.076 (1.022-1.132) |
| 75-79 | 2.973 (2.824-3.129) | 1.580 (1.500-1.663) | 2.276 (2.162-2.396) |
| 80+ | 8.619 (8.187-9.072) | 5.928 (5.631-6.240) | 7.274 (6.909-7.656) |

**Figure S6.**
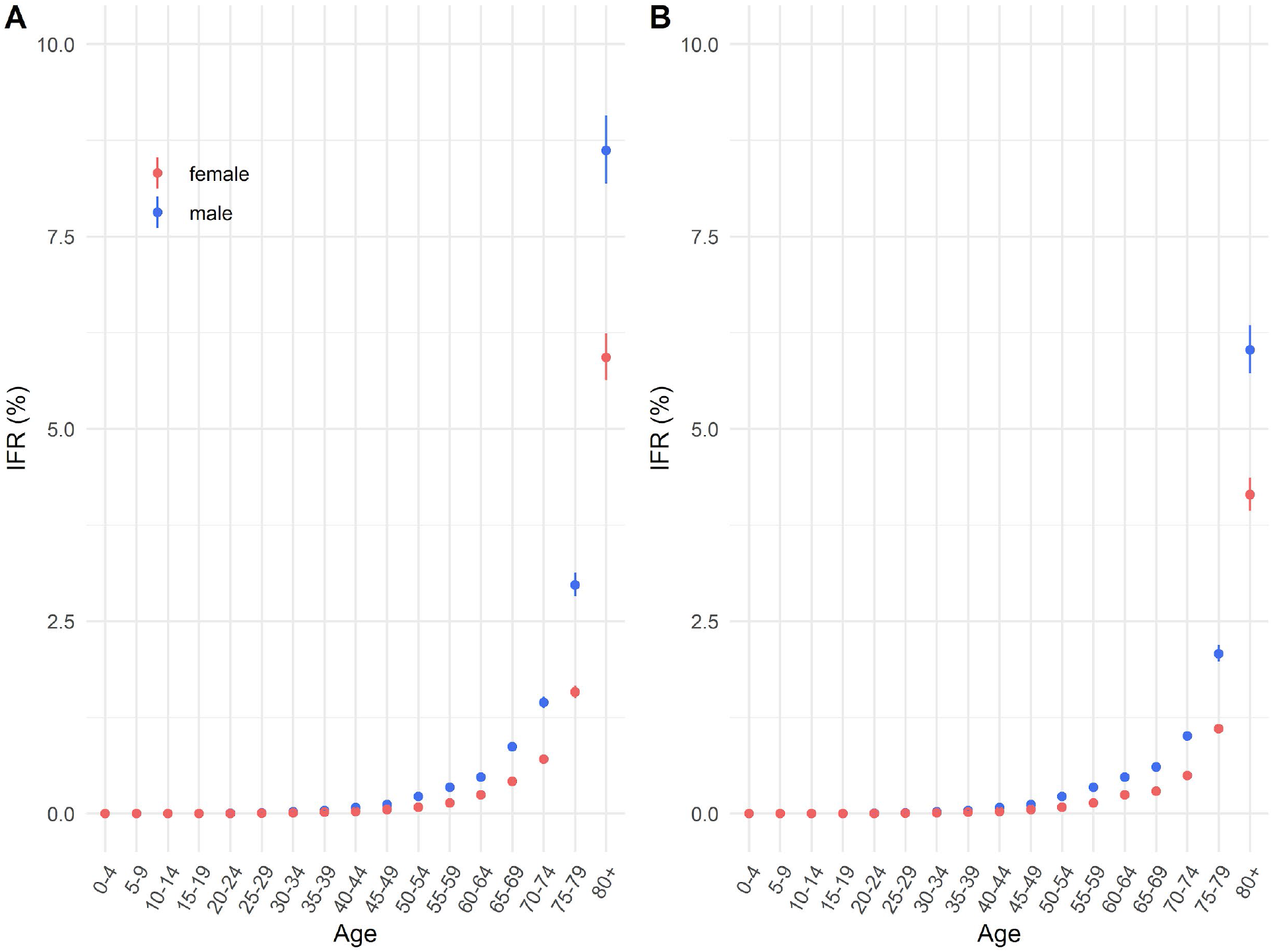
Ensemble age- and sex-specific IFR estimates. (A) Median and 95% credible interval ensemble IFR estimates assuming equal attack rates amongst <65 year olds and a relative attack rate of 0.7 for individuals aged 65+. (B) Median and 95% credible interval ensemble IFR estimates assuming equal attack rates across all ages.

**Supplementary Figure S7.**
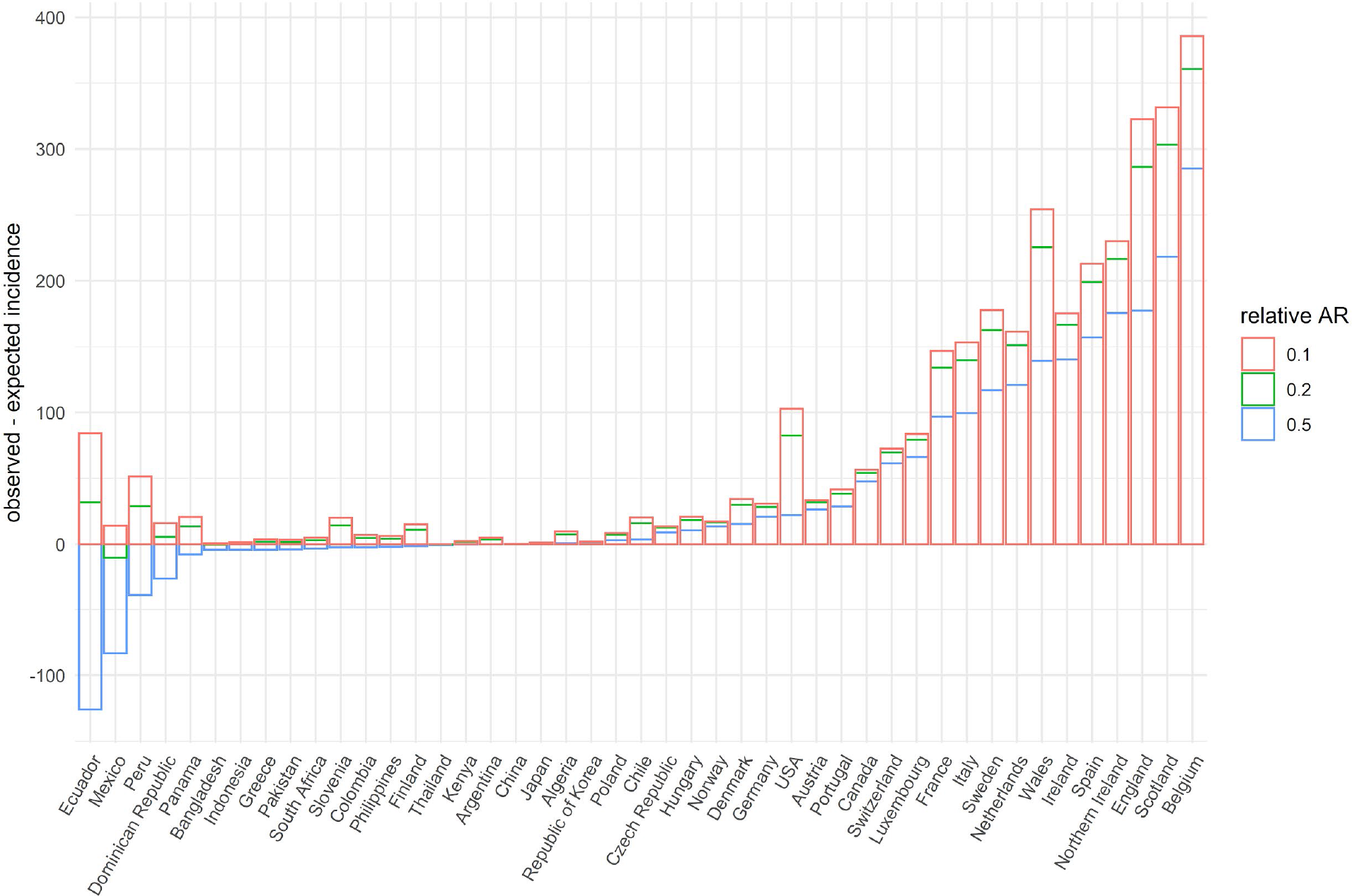
Observed minus expected incidence of deaths aged 60 or 65+, per 100,000 population, by country under different assumptions of the relative infection attack rate (AR) amongst over 65s.

**Figure S8.**
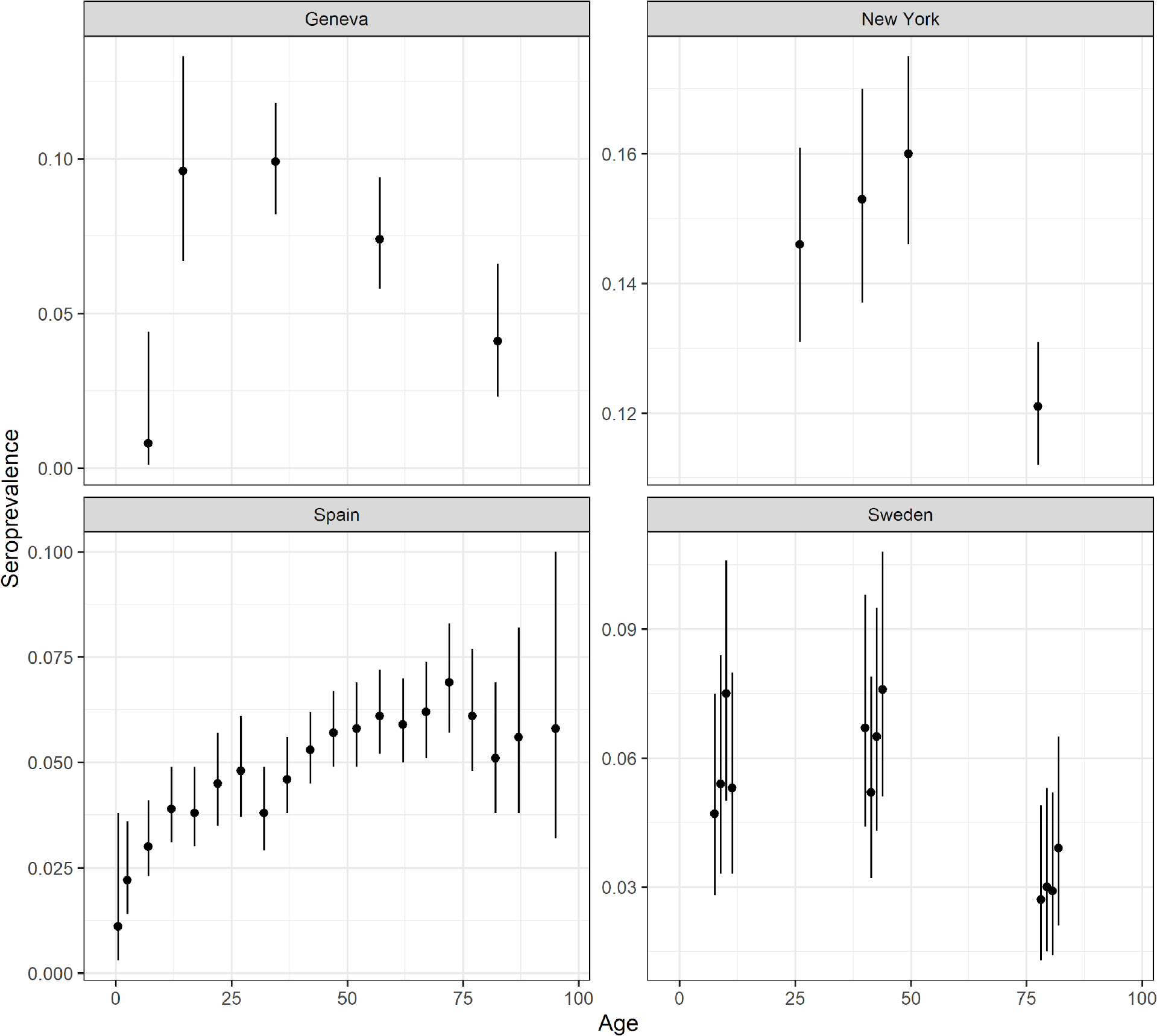
Age-specific seroprevalence data from 4 serostudies^9,13,15,18^, plotted at the age-group mid-points.

